# A Randomized, Placebo-Controlled Trial to Evaluate the Safety and Efficacy of VIR-2482 in Healthy Adults for Prevention of Influenza A Illness (PENINSULA)

**DOI:** 10.1101/2024.04.03.24305209

**Authors:** Susanna K. Tan, Deborah Cebrik, David Plotnik, Maria L. Agostini, Keith Boundy, Christy M. Hebner, Wendy W. Yeh, Phillip S. Pang, Jaynier Moya, Charles Fogarty, Manuchehr Darani, Frederick G. Hayden

## Abstract

**Background:** Influenza A results in significant morbidity and mortality. VIR-2482, an engineered human monoclonal antibody with extended half-life, targets a highly conserved epitope on the stem region of influenza A hemagglutinin, and may protect against seasonal and pandemic influenza.

**Methods:** This double-blind, randomized, placebo-controlled, phase 2 study examined the safety and efficacy of VIR-2482 for seasonal influenza A illness prevention in unvaccinated healthy adults. Participants (N = 2977) were randomized 1:1:1 to receive VIR-2482 450 mg, VIR-2482 1200 mg, or placebo via intramuscular (IM) injection. Primary and secondary efficacy endpoints were the proportions of participants with reverse transcriptase-polymerase chain reaction (RT-PCR)–confirmed influenza A infection and either protocol-defined influenza-like illness (ILI) and Centers for Disease Control and Prevention (CDC)–defined ILI or World Health Organization (WHO)–defined ILI, respectively.

**Results:** VIR-2482 450 mg and 1200 mg prophylaxis did not reduce the risk of protocol-defined ILI with RT-PCR–confirmed influenza A versus placebo (relative risk reduction [RRR], 3.8% [95% CI: −67.3, 44.6] and 15.9% [95% CI: −49.3, 52.3], respectively). At the 1200 mg dose, the RRRs in influenza A illness were 57.2% [95% CI: −2.5, 82.2] using CDC-ILI and 44.1% [95% CI: −50.5, 79.3] using WHO-ILI definitions, respectively. Serum VIR-2482 levels were similar regardless of influenza status; variants with reduced VIR-2482 susceptibility were not detected. Local injection-site reactions were mild and similar across groups.

**Conclusion:** VIR-2482 1200 mg IM was well tolerated but did not significantly prevent protocol-defined ILI. Secondary endpoint analyses suggest this dose may have reduced influenza A illness.

**Trial registration:** ClinicalTrials.gov identifier, NCT05567783

**Key points:** Prophylactic administration of 1200 mg of VIR-2482, an engineered human monoclonal antibody targeting a highly conserved epitope on the stem region of influenza A hemagglutinin, did not significantly reduce risk of influenza-like illness from influenza A virus in healthy adults.

## Introduction

According to the World Health Organization (WHO) approximately 1 billion annual cases of seasonal influenza occur globally. Of these, an estimated 3 to 5 million cases are severe, with up to 650,000 deaths per year [1]. Influenza A and B are both responsible for seasonal epidemics, but influenza A infections account for the majority of hospitalizations and are the only influenza type to cause pandemics [2]. Some patient groups are at high risk of influenza-associated hospitalizations and mortality, including the elderly, immunocompromised hosts, and those with certain comorbidities [3–5].

Currently available seasonal influenza vaccines are influenza strain–specific, incompletely protective due to strain mismatch, and often inadequately immunogenic, especially in high-risk groups like the elderly and immunocompromised [6]. Specifically, vaccine effectiveness against medically attended influenza illness ranged from 19% to 60% in the United States between the 2009 to 2023 influenza seasons [7]. Therefore, we hypothesized that an immunoprophylactic antibody with long half-life and activity against a broad range of influenza A viruses could provide greater protection and obviate the need for updates to match the prevalent circulating strains, especially in individuals at high-risk of complications. Additionally, such a neutralizing antibody could facilitate a rapid response to an influenza A pandemic [8].

VIR-2482 is a neutralizing, engineered human immunoglobulin G (IgG) monoclonal antibody (mAb) that targets a highly conserved epitope on the stem region of the influenza A hemagglutinin (HA) protein [9, 10]. Epidemiologically, HA-stem binding antibodies have been shown to correlate with protection from illness [11] and are the focus of ongoing efforts to create universal influenza vaccines [8, 12].

VIR-2482 and its parent mAb bind to HAs representing all 18 influenza A subtypes and neutralize a broad panel of H1N1 and H3N2 influenza viruses spanning >100 years of antigenic evolution. Administration of VIR-2482 reduced morbidity and mortality from infection by seasonal and zoonotic strains of influenza in mice, ferrets, and macaques [9, 13–15]. VIR-2482 is engineered to include an Fc LS mutation (M428L/N434S) that extends the elimination half-life (T_1/2_) through increased FcRn-mediated antibody recirculation [9].

In a phase 1 study in healthy volunteers (ClinicalTrials.gov Identifier: NCT04033406), VIR-2482 doses of 60 to 1800 mg via intramuscular (IM) injection showed favorable local and systemic tolerability, with few injection-site reactions [10]. Median time to maximum plasma concentration of VIR-2482 ranged from 7.0 to 12.5 days, and median plasma T_1/2_ was 57.1 to 70.6 days across dosing cohorts, indicating the potential for administration once per influenza season.

We undertook this proof-of-concept, phase 2 trial to evaluate the safety, tolerability, and efficacy of VIR-2482 in preventing influenza A illness in healthy adults who had not received a seasonal influenza vaccine. Since vaccination was prohibited, participants were required to be at low risk of developing serious influenza-related complications.

## METHODS

### Study Design

This was a double-blind, randomized, placebo-controlled, dose-ranging study conducted at 53 centers in the United States during the 2022 to 2023 Northern Hemisphere influenza season. The study was conducted in accordance with the principles of the Declaration of Helsinki and the study protocol was approved by the relevant institutional ethics committees (authorizing body: WCG Institutional Review Board). All participants provided written informed consent.

### Participants

Males and nonpregnant/nonlactating females aged 18 to <65 years were eligible to participate if they were in good health as determined from medical history. Key inclusion criteria included body mass index (BMI) 18.0 to 35.0 kg/m^2^ and no clinically significant findings from physical examination, 12-lead electrocardiogram, and laboratory values. Participants were excluded if they had prior or planned receipt of any influenza vaccine for the upcoming season, history or clinical evidence of conditions considered high risk for developing influenza-related complications, or confirmed influenza infection within 3 months of randomization (see Supplementary Methods).

### Antibody Administration and Safety Monitoring

VIR-2482 or saline placebo was administered as 2 separate 4-mL IM injections. Participants were randomized 1:1:1 to VIR-2482 450 mg (1 dose of VIR-2482 and 1 dose of placebo), VIR-2482 1200 mg (2 doses of VIR-2482) or placebo (2 doses of placebo) on Day 1. These doses were selected based on the acceptable safety and tolerability of VIR-2482 doses in the phase 1 study and target serum concentrations estimated from in vitro neutralization data [10]. VIR-2482 was administered as a 150-mg/mL solution. The preferred injection site was the thigh, with the buttock as an alternative.

Participants remained at the clinical study site for ≥2 hours postdose to assess safety, including solicited local tolerability at the injection sites performed at 1-hour postdose. Following discharge, participants completed an electronic diary card from Days 1 to 7 to record injection site and systemic reactions and body temperature. Follow-up visits were conducted at 1, 4, and 12 weeks postdosing and at the end of the influenza season (EOIS).

Adverse events (AEs) were recorded through the end of the study, with severity assessed according to the Toxicity Grading Scale for Healthy Adult and Adolescent Volunteers Enrolled in Preventative Vaccine Clinical Trials [16]. Immunogenicity of VIR-2482 was evaluated according to incidence and titers of antidrug antibodies (ADAs) using validated methods [10].

### Illness Surveillance

Participants completed an electronic diary for influenza-like illness (ILI) symptoms twice per week from Day 1 through EOIS (April 16, 2023). Signs and symptoms included sore throat, cough, sputum production, wheezing, difficulty breathing, temperature >37.8°C, chills, weakness, or myalgia. Participants experiencing any ILI symptoms were requested to visit the study center within 3 days of symptom onset for nasopharyngeal swab (NPS) collection.

Influenza A was confirmed by reverse transcriptase-polymerase chain reaction (RT-PCR) using a sponsor-provided point-of-care device (Xpert-Xpress CoV-2/RSV/Flu plus; Cepheid; Sunnyvale, CA) or a central virology laboratory assay (Respiratory Panel 1; Seegene; Seoul, South Korea) performed at DDL Diagnostic Laboratories (Rijswijk, Netherlands). Participants with ILI were managed according to local standard of care, and symptoms were followed-up daily via telephone through Day 10 following initial presentation. Symptom severity of ILI was documented using the Influenza Intensity and Impact Questionnaire Flu-iiQ™ [17] administered from Day 1 of symptom onset through Day 10.

### Pharmacokinetics

Serum samples for pharmacokinetics (PK) and immunogenicity analyses were collected predose and at the follow-up visits. NPS for PK analysis [10] were collected from all participants at the EOIS visit and at Weeks 1, 4, and 12 from a subset of participants in an optional PK substudy. In participants with ILI, additional samples for PK, ADA, and virologic analyses were collected at the ILI confirmation clinic visit.

### Outcomes

The primary efficacy endpoint was the proportion of participants with RT-PCR positivity for influenza A and protocol-defined ILI. The protocol definition required ≥1 respiratory symptom (sore throat, cough, sputum production, wheezing, or difficulty breathing) and ≥1 systemic sign or symptom (temperature >37.8°C, chills, weakness, or myalgia). Prespecified secondary efficacy endpoints were the proportions of participants with RT-PCR–confirmed influenza A and ILI as defined by the Centers for Disease Control and Prevention (CDC; temperature >37.8°C with sore throat or cough and RT-PCR confirmation) and WHO (temperature >38°C with cough and RT-PCR confirmation).

Exploratory endpoints included the severity and duration, as the time of onset of first symptom through when all respiratory symptoms were resolved for ≥24 hours, of participant-reported ILI signs and symptoms with the Flu-iiQ™ instrument [17]. Virologic analyses on NPS samples determined the proportion of participants with RT-PCR–confirmed influenza A by virus subtype and assessed emergence of virus variants with reduced susceptibility to VIR-2482. Next-generation sequencing of the influenza A HA gene was performed at DDL Diagnostic Laboratories (Rijswijk, Netherlands). Virus successfully cultured in MDCK cells was assessed for reduced susceptibility using an in vitro neutralization assay at Viroclinics Biosciences (Rotterdam, Netherlands) [18]. Sequence and phenotypic results were compared with respective subtype vaccine reference strains (H1N1: A/Victoria/2750/2019; H3N2: A/Darwin/9/2021 and A/Darwin/6/2021, respectively).

### Statistical Analysis

A sample size of approximately 3000 participants was needed to provide approximately 80% power to detect VIR-2482 efficacy (a true relative risk reduction [RRR] of 70%) of 1 VIR-2482 dose level compared with placebo, assuming an attack rate of ≥2.25% in the placebo group [19, 20]. Efficacy and safety were analyzed in all randomized participants receiving any amount of study drug or placebo. Efficacy was assessed based on the study intervention to which participants were randomly assigned, and safety was assessed based on the intervention received. Details on primary and secondary estimands and statistical analyses are provided in the Supplementary Methods.

A modified Poisson regression with robust variance was used as the efficacy analysis model to estimate the relative risk of the incidence of protocol-/CDC-/WHO-defined ILI with RT-PCR–confirmed influenza A in each VIR-2482 group compared with the placebo group, with treatment group as the only factor in each model. A hierarchical fixed sequence testing procedure was used to control the overall type I error as outlined in the Supplementary Methods. Formal comparisons precluded by the hierarchical testing strategy were considered descriptive.

## Results

### Participants and Disposition

Between October 13, 2022, and January 6, 2023, participants (N = 2977) were randomized, and 2956 (99.3%) received study intervention (**Figure 1**). Across treatment groups, mean age was 39.5 years and 54% were female (**Table 1**). Study drug or placebo was administered in the thigh in approximately 78% of participants. Through the end of study, discontinuation rates were <5% and comparable across treatment groups.

**Figure 1.**
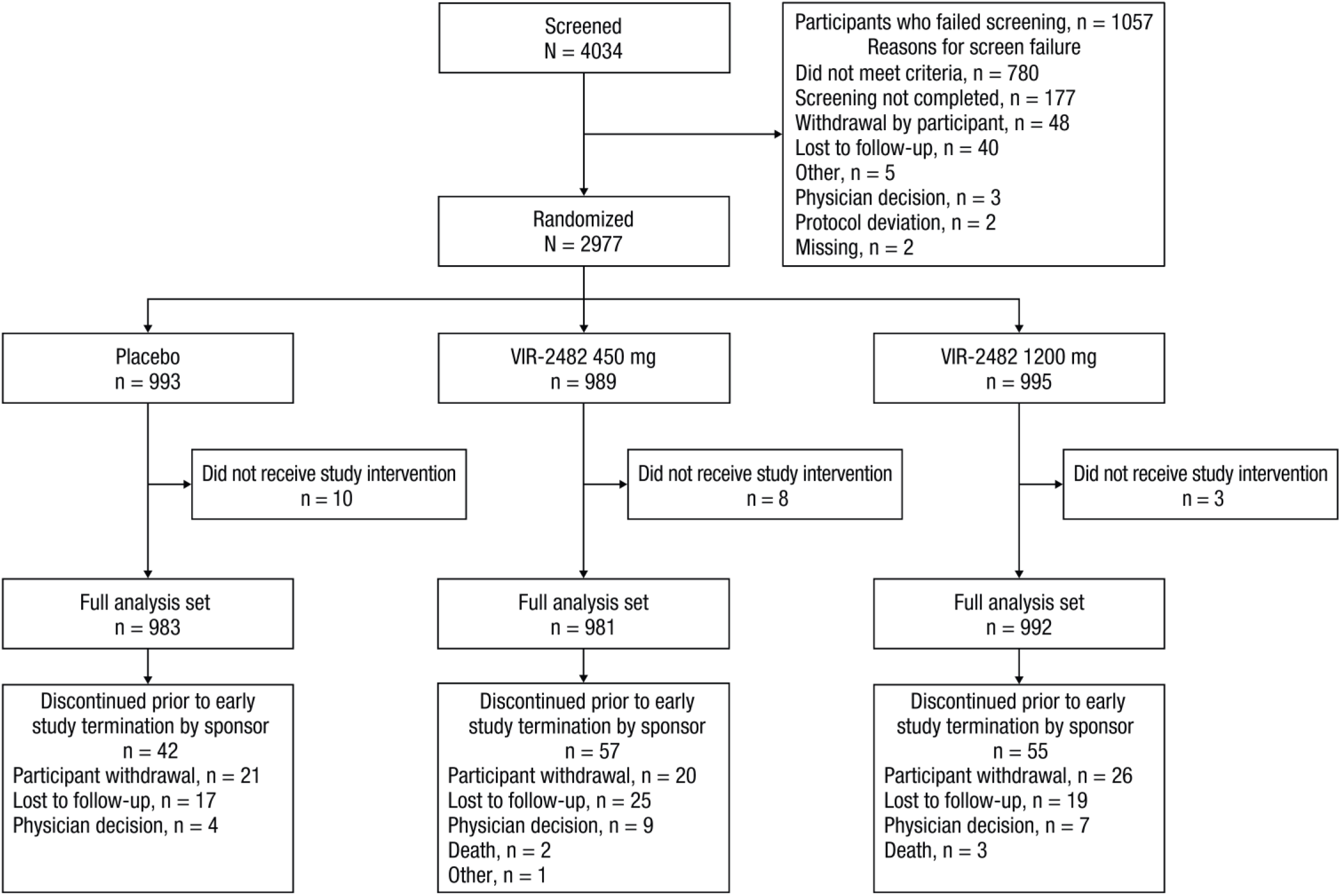
Participant Disposition. Based on efficacy findings, the study was terminated early and follow-up into the second influenza season did not take place.

**Table 1.** Baseline Characteristics (Full Analysis Set)

|  | Placebo (n =<br>983) | VIR-2482 450<br>mg (n = 981) | VIR-2482<br>1200 mg (n =<br>992) | Total<br>(N = 2956) |
| --- | --- | --- | --- | --- |
| Age at randomization, years |  |  |  |  |
| Mean (SD) | 39.7 (12.3) | 39.2 (12.5) | 39.6 (11.7) | 39.5 (12.2) |
| Median (range) | 39.0 (18-64) | 38.0 (18-64) | 39.0 (18-64) | 39.0 (18-64) |
| Sex, no. (%) |  |  |  |  |
| Male | 453 (46.1) | 436 (44.4) | 469 (47.3) | 1358 (45.9) |
| Female | 530 (53.9) | 545 (55.6) | 523 (52.7) | 1598 (54.1) |
| Race, no. (%) |  |  |  |  |
| American Indian or<br>Alaskan native | 7 (0.7) | 7 (0.7) | 7 (0.7) | 21 (0.7) |
| Asian | 26 (2.6) | 26 (2.7) | 29 (2.9) | 81 (2.7) |
| Black or African<br>American | 166 (16.9) | 154 (15.7) | 176 (17.7) | 496 (16.8) |
| Native Hawaiian or<br>other Pacific Islander | 4 (0.4) | 5 (0.5) | 4 (0.4) | 13 (0.4) |
| White | 758 (77.1) | 752 (76.7) | 747 (75.3) | 2257 (76.4) |
| Multiracial | 6 (0.6) | 17 (1.7) | 11 (1.1) | 34 (1.2) |
| Not<br>reported/unknown | 16 (1.6) | 20 (2.0) | 18 (1.8) | 54 (1.8) |
| Ethnicity, no. (%) |  |  |  |  |
| Hispanic or Latino | 357 (36.3) | 370 (37.7) | 370 (37.3) | 1097 (37.1) |
| Not Hispanic or<br>Latino | 620 (63.1) | 607 (61.9) | 621 (62.6) | 1848 (62.5) |
| Not<br>reported/unknown | 6 (0.6) | 4 (0.4) | 1 (0.1) | 11 (0.4) |
| Body mass index, kg/m<sup>2</sup> |  |  |  |  |
| Mean (SD) | 27.2 (4.1) | 27.3 (4.1) | 27.4 (4.2) | 27.3 (4.1) |
| Median (range) | 27.2 (18.0-35.0) | 27.5 (18.0-36.6) | 27.7 (18.0-35.1) | 27.5 (18.0-36.6) |
| Injection site location, no. (%) |  |  |  |  |
| Thigh | 762 (77.5) | 757 (77.2) | 783 (78.9) | 2302 (77.9) |
| Buttock | 219 (22.3) | 223 (22.7) | 207 (20.9) | 649 (22.0) |

### Efficacy Outcomes

The primary endpoint of protocol-defined ILI with RT-PCR confirmed influenza A was observed in similar numbers of participants across groups (**Table 2**). A nonsignificant RRR of 15.9% (95% CI: −49.3, 52.6) comparing VIR-2482 1200 mg versus placebo was observed. Predefined subgroup analyses between these 2 groups showed RRRs consistent with that of the overall population (**Supplementary Figure 1**). In the VIR-2482 1200 mg group, RRRs versus placebo for CDC- and WHO-defined ILI with RT-PCR–confirmed influenza A were 57.2% (95% CI: - 2.5, 82.2) and 44.1% (95% CI: −50.5, 79.3), respectively (**Table 2**).

**Table 2.** Occurrence of Influenza-Like Illness With RT-PCR Confirmed Influenza A Through End of Influenza Season (Full Analysis Set)

|  | Placebo<br>(n = 983) | VIR-2482 450 mg<br>(n = 981) | VIR-2482 1200<br>mg (n = 992) |
| --- | --- | --- | --- |
| Primary endpoint |  |  |  |
| No. (%) participants with<br>protocol-defined ILI<sup>a</sup> | 25 (2.54) | 24 (2.45) | 21 (2.12) |
| Relative risk reduction vs<br>placebo, % | - | 3.78 | 15.85 |
| 95% CI (%)<sup>b</sup> | - | -67.23, 44.63 | -49.27, 52.56 |
| P-value | - | - | 0.56 |
| Secondary endpoints |  |  |  |
| No. (%) participants with<br>CDC-defined ILI<sup>a</sup> | 17 (1.73) | 15 (1.53) | 7 (0.71) |
| Relative risk reduction vs<br>placebo, % | - | 11.45 | 57.23 |
| 95% CI (%)<sup>b</sup> | - | -76.25, 55.51 | -2.51, 82.15 |
| No. (%) participants with<br>WHO-defined ILI<sup>a</sup> | 11 (1.12) | 12 (1.22) | 6 (0.6) |
| Relative risk reduction vs<br>placebo, % | - | -9.80 | 44.13 |
| 95% CI (%)<sup>b</sup> | - | -147.41, 51.27 | -50.49, 79.26 |

CDC, Centers for Disease Control and Prevention; ILI, influenza-like illness; RT-PCR, reverse transcriptase-polymerase chain reaction; WHO, World Health Organization.
<sup>a</sup>Participants with multiple occurrences of protocol-, CDC-, or WHO-defined illness with RT- PCR–confirmed influenza A were counted once.
<sup>b</sup>The 95% CI for the primary endpoint analysis of 450 mg versus placebo and for analyses of secondary endpoints are nominal, and not adjusted for multiplicity.

To investigate the impact of pre-existing infections or infections that may have been acquired prior to VIR-2482 achieving adequate tissue distribution, we conducted posthoc analyses excluding 10 cases of protocol-defined ILI, 5 cases of CDC-defined ILI, and 4 cases of WHO-defined ILI that occurred within 7 days following study drug administration. Numerically higher RRRs of ILI were observed in the VIR-2482 1200 mg group (protocol-defined: 34.0% [95% CI: −25.7, 65.3]; CDC-defined: 64.9% [95% CI: 3.9, 87.2]; WHO-defined: 42.9% [95% CI: −69.7, 80.8]; **Supplementary Table 1**).

Most participants with protocol-defined ILI reported moderate to severe symptoms. Symptom severity of ILI measured by FluiiQ™ was similar across groups (**Supplemental Table 2**). Participants who received VIR-2482 1200 mg had a numerically shorter time to resolution of protocol-defined ILI than participants who received placebo (median: 85.8 h [interquartile range (IQR): 57.7-137.2] vs 111.9 h [IQR: 72.1-130.4]). No influenza-related hospitalizations or deaths were reported.

### Safety

Treatment-emergent AEs (TEAEs) were reported in 62% to 65% of participants across study groups (**Table 3**). The most frequently reported TEAEs (>5% of participants in any group) were upper respiratory tract infection, COVID-19, oropharyngeal pain, cough, viral infection, and myalgia. Treatment-related TEAEs were reported in ≤2% of participants and were similarly distributed across groups. Serious TEAEs were uncommon (≤1% across groups), and none were considered related to study treatment. Five deaths occurred during the study, and none were considered related to treatment.

**Table 3.** Occurrences of TEAEs Through the End of the Study (Safety Set)

|  | Placebo<br>(n = 983)<br>n (%) | VIR-2482 450 mg<br>(n = 981)<br>n (%) | VIR-2482 1200 mg<br>(n = 992)<br>n (%) |
| --- | --- | --- | --- |
| Any TEAE | 639 (65.0) | 636 (64.8) | 613 (61.8) |
| TEAEs by maximum toxicity grade |  |  |  |
| Grade 1 | 441 (44.9) | 449 (45.8) | 406 (40.9) |
| Grade 2 | 181 (18.4) | 174 (17.7) | 190 (19.2) |
| Grade 3 | 14 (1.4) | 10 (1.0) | 12 (1.2) |
| Grade 4 | 3 (0.3) | 3 (0.3) | 5 (0.5) |
| Study treatment–related TEAEs | 15 (1.5) | 20 (2.0) | 17 (1.7) |
| TEAEs leading to study treatment discontinuation or interruption | 0 | 0 | 0 |
| Serious TEAEs | 9 (0.9) | 10 (1.0) | 13 (1.3) |
| Study treatment–related serious TEAEs | 0 | 0 | 0 |
| TEAEs leading to death <sup>a</sup> | 0 | 2 (0.2) | 3 (0.3) |
| TEAEs occurring in >5% of participants in any treatment group |  |  |  |
| Upper respiratory tract infection | 248 (25.2) | 243 (24.8) | 245 (24.7) |
| COVID-19 | 106 (10.8) | 93 (9.5) | 92 (9.3) |
| Oropharyngeal pain | 75 (7.6) | 95 (9.7) | 87 (8.8) |
| Cough | 72 (7.3) | 88 (9.0) | 84 (8.5) |
| Viral infection | 70 (7.1) | 56 (5.7) | 48 (4.8) |
| Myalgia | 38 (3.9) | 51 (5.2) | 36 (3.6) |

TEAE, treatment-emergent adverse event.
A participant with multiple occurrences of the same TEAE was counted only once at the maximum toxicity grade or strongest relationship to study intervention.
<sup>a</sup>Reasons for death were accidental drug overdose (nontreatment intervention) and suicide (both VIR-2482 450 mg), cardiac arrhythmia, cardiorespiratory arrest, and intentional multidrug overdose (nontreatment intervention; all VIR-2482 1200 mg). None of the deaths were considered related to study treatment.

Incidences of solicited injection-site reactions at 1-hour postdose were <10% in all 3 groups (**Supplementary Table 3**). Grade 1 (mild) injection-site pain was the most common self-reported injection-site reaction (placebo, 5.6%; VIR-2482 450 mg, 6.1%; VIR-2482 1200 mg, 8.4%) during the on-site assessment. Mild injection-site pain was also the most common injection-site reaction reported on the electronic diary card during Days 1 to 7 (placebo, 22.6%; VIR-2482 450 mg, 24.5%; VIR-2482 1200 mg, 23.9%; **Supplementary Table 4**). The most common participant-reported systemic reactions included mild myalgia (12.3%-14.9%), mild headache (12.5%-14.0%), and mild malaise (9.8%-10.3%; **Supplementary Table 5**). No treatment-emergent anaphylaxis events were reported. Incidences of treatment-emergent clinical laboratory and vital signs abnormalities were similar across groups.

### Virologic Analyses

Among participants with protocol-defined ILI with RT-PCR–confirmed influenza A, H3 subtype virus was detected in 56% to 72% and H1 subtype in 16% to 36% of illnesses across study groups; 8% to 19.1% of illnesses had an unknown influenza A subtype (**Supplementary Table 6**).

Sequence and phenotypic data availability are described in **Supplementary Table 7**. Influenza A virus isolates, including the small number where a VIR-2482 epitope amino acid substitution was detected by sequence analysis, retained susceptibility to VIR-2482 with fold changes in half maximal effective concentration (EC_50_) ranging from 0.23 to 2.55 compared with the respective subtype vaccine reference strain in vitro.

### Pharmacokinetics

Following VIR-2482 1200 mg, median serum T_max_ was 6.95 days, geometric mean serum C_max_ was 111 µg/mL (CV% = 52.1%), and geometric mean serum concentration at EOIS was 18.5 µg/mL (CV% = 98.8). Median T_1/2_ was 54.7 to 55.4 days across VIR-2482 doses. Compared with gluteal administration, thigh injection resulted in 79.9% higher C_max_ and 43.8% higher area under the curve (AUC) for the 450 mg dose and 97.5% higher C_max_ and 55.6% higher AUC for the 1200 mg dose (**Supplementary Table 8**).

Serum and NPS concentration-time data through 180 days postdose are displayed in **Figure 2** and NPS:serum ratios are displayed in **Supplemental Figure S2**. The NPS concentrations of VIR-2482 averaged 2.1% to 7.8% of within-subject serum concentrations across doses and time points. VIR-2482 recipients who experienced breakthrough influenza A illness had serum levels that were comparable to those who did not (**Supplemental Table 9**).

**Figure 2.**
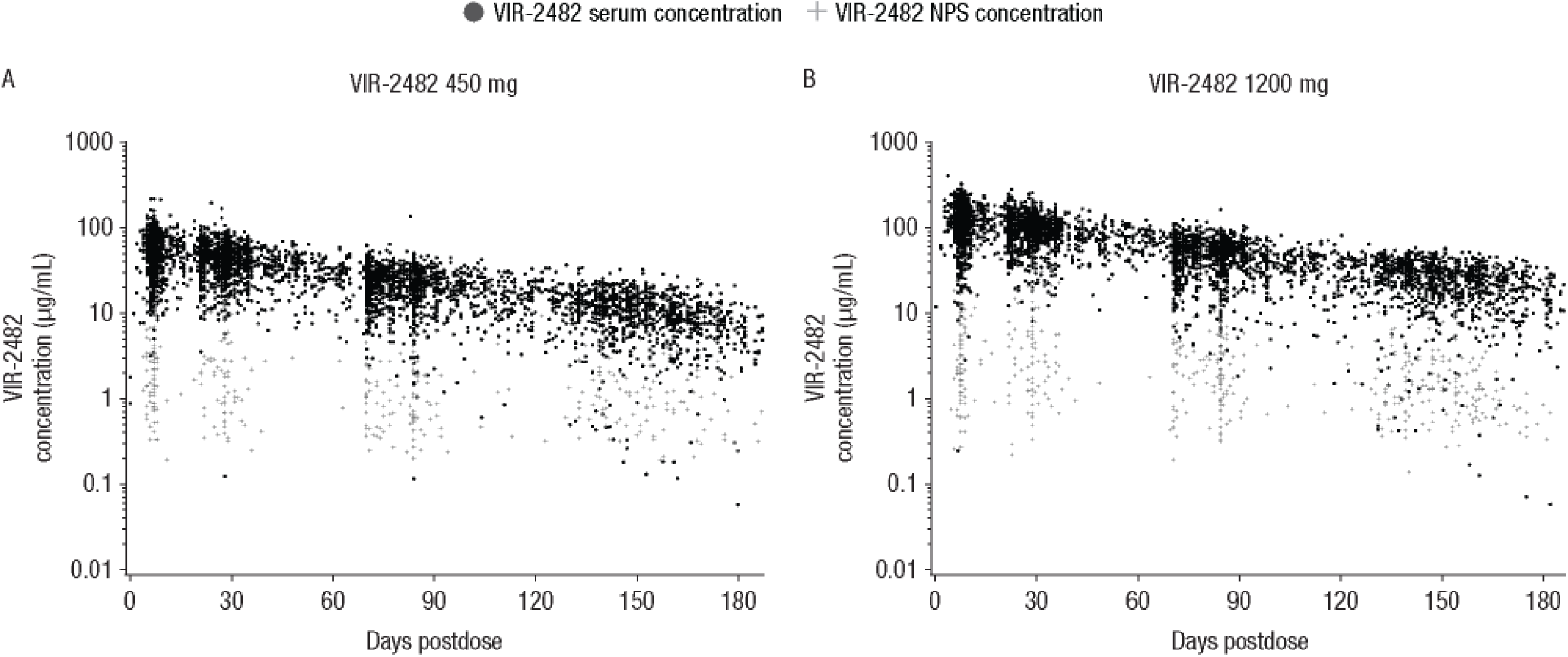
Pharmacokinetic Profiles of VIR-2482 Following Intramuscular Administration of (A) 450-mg and (B) 1200 mg Doses. NPS, nasopharyngeal swab.

### Immunogenicity

The incidence of ADA was 5.4% (53/980) in the 450 mg group and 5.1% (51/992) in the 1200 mg group. A >4-fold increase in ADA titer relative to baseline was observed in 1 participant receiving VIR-2482 1200 mg, while the titer of all other pre-existing ADAs was unaffected by treatment. Further, ADAs were not correlated with AEs, injection-site reactions, or aberrant PK in any participant (data not shown).

## Discussion

Neutralizing mAbs have demonstrated the ability to prevent symptomatic respiratory diseases, such as COVID-19 in adults [21] and respiratory syncytial virus (RSV) in infants [22]. Broadly neutralizing HA-stem binding antibodies are recognized as potential pandemic influenza countermeasures [13, 23], and studies of such antibodies have shown therapeutic efficacy in experimentally induced human influenza infections [24, 25] but have demonstrated inconsistent efficacy in treating uncomplicated seasonal influenza [26–28]. The current trial found that prophylactic IM VIR-2482 administration was generally well tolerated, but did not significantly protect against protocol-defined ILI with RT-PCR–confirmed influenza A compared with placebo. Several explanations could account for the lack of significant clinical efficacy observed in this study.

Two potential contributory factors were the early onset of influenza activity in the 2022 to 2023 influenza season [29] that coincided with study enrollment and dosing and the known lag between maximal serum and tissue concentrations following IM dosing. When RT-PCR– confirmed illnesses that occurred <7 days after dosing were excluded, posthoc analyses showed greater reductions in ILI in VIR-2482 1200 mg recipients, including 65% RRR of the CDC-defined ILI cases in the 1200 mg group compared with placebo. These results suggest that early ILIs could have been due to either incubating infections prior to VIR-2482 dosing or infections acquired during the window prior to adequate tissue distribution. In addition, we noted that participants who received VIR-2482 1200 mg had a numerically shorter time to resolution of protocol-defined ILI than participants who received placebo, suggesting clinical activity at this dose.

The apparent dose-response observed in those who received VIR-2482 1200 mg versus 450 mg suggests the possibility that the onset and durability of protection was potentially limited by the drug levels achieved in relevant tissue compartments. The VIR-2482 1200 mg dose selection was based on modelling using in vitro EC_90_ data that estimated it would provide antiviral activity for >8 months [10]. Additional PK-pharmacodynamics analyses are needed to better understand the tissue distribution of VIR-2482 and the optimal drug levels needed in the respiratory tract.

The serum and nasopharyngeal exposures of VIR-2482 observed in this study were comparable to those reported in the phase 1 study of healthy volunteers [10]. Also, the overall incidence of ADAs was low and not correlated with reduced serum concentrations or incidence of infections. Consequently, dosing errors, manufacturing concerns, or aberrant PK are unlikely explanations for trial failure. Viral resistance was also unlikely to explain the lack of efficacy, since viruses cultured from participants with influenza A infections retained susceptibility to VIR-2482 compared with respective subtype reference viruses in vitro.

One hypothesis that may account for these results is that the antiviral activities of HA-stem antibodies observed in vitro or in in vivo animal models do not translate into clinically meaningful benefit when administered to unvaccinated healthy adults for prophylaxis of symptomatic infection. The HA-stem antibodies that block fusion of the endocytosed virion within cells may not be as effective at preventing symptomatic infection compared with the head-binding HA antibodies typically induced by seasonal vaccines that block receptor binding.

An additional hypothesis is that VIR-2482 has clinically meaningful activity, but the selected endpoints, dose, study population, or timing did not enable a positive trial result. Unvaccinated healthy adults were selected as the trial population in this study to better understand the tolerability profile and clinical activity of VIR-2482 without prior vaccination. The prespecified CDC-defined secondary endpoint, which requires a temperature >37.8°C, demonstrated numerically a 57.2% RRR in symptomatic RT-PCR–confirmed infection in VIR-2482 1200 mg recipients versus the 15.9% RRR observed for the primary endpoint. This raises the possibility that VIR-2482 could be more effective in preventing severe or systemic influenza illness in a more vulnerable patient population than unvaccinated healthy adults. Indeed, this concept has been demonstrated with mAbs targeting RSV in infants, where the primary endpoint is not mild symptomatic illness, but medically attended lower respiratory tract infection [22]. COVID-19 vaccines and mAbs have also demonstrated effectiveness against severe disease but are less effective in protecting against mild illness [30–32].

In conclusion, although VIR-2482 was well tolerated, it did not significantly prevent influenza A illness in healthy unimmunized adults at IM doses up to 1200 mg. There remains a clinical need for long-duration prophylactic interventions that provide protection against the morbidity and mortality of influenza in individuals at high-risk of complications. The findings from this trial provide valuable insights to support the future development of broad spectrum mAbs for the prevention of influenza illness.

## Data Availability

Relevant data can be found within the manuscript and supplemental data.

## Acknowledgements

The authors thank all study participants, investigators, site personnel, and operational staff. The authors also thank Jeanne McKeon, PhD, of Lumanity Scientific Inc., for medical writing and editorial support, which was funded by Vir Biotechnology, Inc.

## Author Contributions

Susanna K. Tan, Deborah Cebrik, David Plotnik, Maria L. Agostini, Keith Boundy, Christy M. Hebner, Wendy W. Yeh, Philip S. Pang, and Frederick G. Hayden conceptualized and designed the study. Jaynier Moya, Charles Fogarty, and Manuchehr Darani acquired and analyzed data. Deborah Cebrik conducted the statistical analyses. All authors interpreted data, drafted the manuscript, and critically reviewed and revised the manuscript for important intellectual content. All authors had full access to all the data in the study, take responsibility for the accuracy of the analysis, and had authority over manuscript preparation and the decision to submit the manuscript for publication.

## Data Sharing Statement

Relevant data can be found within the manuscript and supplemental data.

## Supplementary Methods

### Participant Exclusion Criteria

Participants were excluded from this study if they had an established diagnosis of a condition considered high risk for developing influenza-related complications. Such conditions are listed below:

- Asthma.
- Neurologic and neurodevelopmental conditions (such as cerebral palsy, epilepsy [seizure disorders], stroke, intellectual disability, moderate to severe developmental delay, muscular dystrophy, or spinal cord injury).
- Chronic lung disease (such as chronic obstructive pulmonary disease, emphysema, and cystic fibrosis).
- Heart disease (such as congenital heart disease, congestive heart failure, and coronary artery disease).
- Blood disorders (such as sickle cell disease and thalassemia).
- Endocrine disorders (such as diabetes and adrenal insufficiency).
- Chronic kidney disease.
- Chronic liver disease.
- Metabolic disorders (such as inherited metabolic disorders and mitochondrial disorders).
- Weakened immune system due to disease or medication (such as individuals with HIV or AIDS, or cancer, or those taking chronic steroids).
- People younger than 19 years of age who are receiving long-term aspirin therapy.

### Statistical Analysis

The primary estimand was defined as the relative risk of VIR-2482 versus placebo of protocol-defined ILI with central virology laboratory or point-of-care RT-PCR–confirmed influenza A in the absence of RT-PCR–confirmed coinfection with influenza B, respiratory syncytial virus, and/or severe acute respiratory syndrome coronavirus-2 through EOIS in the full analysis set.

The treatment policy strategy was used for the handling of intercurrent events of receipt of any nonstudy influenza antiviral and receipt of an authorized influenza vaccine. Participants who died due to influenza A were to be considered to have met the primary endpoint. Participants who died for reasons other than influenza A illness were handled using the while-alive approach, where data up until the time of death were included in the analysis. The secondary estimands were defined similarly to the primary estimand, with all attributes being the same other than the endpoints (CDC- and WHO-defined ILI with central virology laboratory or point-of-care RT-PCR–confirmed influenza A in the absence of RT-PCR–confirmed coinfection with influenza B, respiratory syncytial virus, and/or severe acute respiratory syndrome coronavirus-2 through EOIS).

VIR-2482 efficacy was calculated as the relative risk reduction in the occurrence of protocol-/CDC-/WHO-defined ILI with RT-PCR–confirmed influenza A in each VIR-2482 group compared with that in the placebo group, or 100% × (1 – the relative risk), with the result expressed as a percentage, with corresponding 95% CI. Participants with multiple occurrences of protocol-/CDC-/WHO-defined ILI with RT-PCR–confirmed influenza A were counted once at the earliest occurrence. Missing endpoint data were imputed with the observed incidence of protocol-/CDC-/WHO-defined ILI with RT-PCR–confirmed influenza A in the placebo group by means of multiple imputation. Exploratory analyses were conducted in predefined subgroups using the same methodology as the primary efficacy endpoint.

### Hierarchical Testing

For the primary estimand, the comparison of VIR-2482 450 mg versus placebo was tested only if the comparison of VIR-2482 1200 mg versus placebo was statistically significant at the 0.05 level (2-sided). Statistical testing was to continue for each comparison for the secondary estimands only if the comparison of VIR-2482 450 mg versus placebo for the primary estimand was tested and statistically significant at the 0.05 level (2-sided).

---

**Supplementary Table 1.** Posthoc: Participants With RT-PCR Confirmed Influenza A ILI Through EOIS, Excluding RT-PCR Confirmed Influenza A ILI With Onset in the First 7 Days Following Study Intervention Administration (Full Analysis Set)

|  | Placebo (n = 983) | VIR-2482 450 mg (n = 981) | VIR-2482 1200 mg (n = 992) |
| --- | --- | --- | --- |
| No. (%) participants with protocol-defined ILI with RT-PCR–confirmed influenza A<sup>a</sup> | 23 (2.3) | 22 (2.2) | 15 (1.5) |
| Relative risk reduction (95% CI) for VIR-2482 vs placebo<sup>b</sup> |  | 4.14 (-70.82, 46.21) | 34.01 (-25.67, 65.34) |
| No. (%) participants with CDC-defined ILI with RT-PCR confirmed influenza A<sup>a</sup> | 15 (1.53) | 14 (1.43) | 5 (0.50) |
| Relative risk reduction (95% CI) for VIR-2482 vs placebo<sup>b</sup> |  | 6.52 (-92.60, 54.63) | 64.88 (3.87, 87.17) |
| No. (%) participants with WHO-defined ILI with RT-PCR–confirmed influenza A<sup>a</sup> | 9 (0.92) | 11 (1.12) | 5 (0.50) |
| Relative risk reduction (95% CI) for<br>VIR-2482 vs placebo<sup>b</sup> |  | -22.86 (-194.88, 48.81) | 42.86 (-69.69, 80.76) |

CDC, Centers for Disease Control and Prevention; EOIS, end of influenza season; ILI, influenza-like illness; RT-PCR, reverse transcriptase-polymerase chain reaction; WHO, World Health Organization.
<sup>a</sup>Participants with multiple occurrences of protocol-defined, CDC-defined, or WHO-defined illness with RT-PCR–confirmed influenza A were counted once.
<sup>b</sup>The 95% CIs are nominal, and not adjusted for multiplicity.

**Supplementary Table 2.** Symptoms Among Participants With Protocol-Defined ILI With RT-PCR**–**Confirmed Influenza A Without Concomitant Coinfection Based on Flu-IIQ, Overall and by Maximum Severity.

| Symptoms, no. (%) | Placebo<br>(n = 983) | VIR-2482 450 mg<br>(n = 981) | VIR-2482 1200 mg<br>(n = 992) |
| --- | --- | --- | --- |
| Participants with any symptoms | 25 (100) | 23 (95.8) | 21 (100) |
| Mild | 0 | 1 (4.2) | 2 (9.5) |
| Moderate | 15 (60.0) | 9 (37.5) | 7 (33.3) |
| Severe | 10 (40.0) | 13 (54.2) | 12 (57.1) |
| Cough | 25 (100) | 23 (95.8) | 20 (95.2) |
| Mild | 5 (20.0) | 7 (29.2) | 7 (33.3) |
| Moderate | 16 (64.0) | 10 (41.2) | 10 (47.6) |
| Severe | 4 (16.0) | 6 (25.0) | 3 (14.3) |
| Sore Throat | 23 (92.0) | 18 (75.0) | 20 (95.2) |
| Mild | 14 (56.0) | 5 (20.8) | 11 (52.4) |
| Moderate | 7 (28.0) | 9 (37.5) | 7 (33.3) |
| Severe | 2 (8.0) | 4 (16.7) | 2 (9.5) |
| Headache | 21 (84.0) | 19 (79.2) | 21 (100) |
| Mild | 5 (20.0) | 9 (37.5) | 5 (23.8) |
| Moderate | 11 (44.0) | 5 (20.8) | 11 (52.4) |
| Severe | 5 (20.0) | 5 (20.8) | 5 (23.8) |
| Nasal congestion | 23 (92.0) | 22 (91.7) | 18 (85.7) |
| Mild | 6 (24.0) | 4 (16.7) | 4 (19.1) |
| Moderate | 13 (52.0) | 14 (58.3) | 9 (42.9) |
| Severe | 4 (16.0) | 4 (16.7) | 5 (23.8) |
| Feeling feverish | 21 (84.0) | 18 (75.0) | 18 (85.7) |
| Mild | 5 (20.0) | 7 (29.2) | 6 (28.6) |
| Moderate | 11 (44.0) | 6 (25.0) | 6 (28.6) |
| Severe | 5 (20.0) | 5 (20.8) | 6 (28.6) |
| Body aches and pains | 22 (88.0) | 19 (79.2) | 20 (95.2) |
| Mild | 5 (20.0) | 9 (37.5) | 8 (38.1) |
| Moderate | 11 (44.0) | 4 (16.7) | 6 (28.6) |
| Severe | 6 (24.0) | 6 (25.0) | 6 (28.6) |
| Fatigue | 25 (100) | 22 (91.7) | 19 (90.5) |
| Mild | 8 (32.0) | 7 (29.2) | 3 (14.3) |
| Moderate | 11 (44.0) | 12 (50.0) | 11 (52.4) |
| Severe | 6 (24.0) | 3 (12.5) | 5 (23.8) |
| Neck pain | 19 (76.0) | 15 (62.5) | 14 (66.7) |
| Mild | 5 (20.0) | 10 (41.7) | 5 (23.8) |
| Moderate | 11 (44.0) | 3 (12.5) | 7 (33.3) |
| Severe | 3 (12.0) | 2 (8.3) | 2 (9.5) |
| Interrupted sleep | 23 (92.0) | 17 (70.8) | 19 (90.5) |
| Mild | 5 (20.0) | 6 (25.0) | 5 (23.8) |
| Moderate | 10 (40.0) | 5 (20.8) | 8 (38.1) |
| Severe | 8 (32.0) | 6 (25.0) | 6 (28.6) |
| Loss of appetite | 23 (92.0) | 18 (75.0) | 18 (85.7) |
| Mild | 11 (44.0) | 9 (37.5) | 9 (42.9) |
| Moderate | 10 (40.0) | 6 (25.0) | 5 (23.8) |
| Severe | 2 (8.0) | 3 (12.5) | 4 (19.1) |

ILI, influenza-like illness; Flu-IIQ, Influenza Intensity and Impact Questionnaire; RT-PCR, reverse transcriptase-polymerase chain reaction.

**Supplementary Table 3.** Solicited Injection-Site Reactions at 1 Hour Postdose (Safety Set)

|  | Placebo (n = 983)<br>n (%) | VIR-2482 450 mg<br>(n = 981)<br>n (%) | VIR-2482 1200<br>mg (n = 992)<br>n (%) |
| --- | --- | --- | --- |
| Any injection-site reaction | 65 (6.6) | 81 (8.3) | 93 (9.4) |
| Grade 1 | 63 (6.4) | 79 (8.1) | 90 (9.1) |
| Grade 2 | 2 (0.2) | 2 (0.2) | 3 (0.3) |
| Pain | 57 (5.8) | 60 (6.1) | 83 (8.4) |
| Grade 1 | 55 (5.6) | 60 (6.1) | 83 (8.4) |
| Grade 2 | 2 (0.2) | 0 | 0 |
| Swelling | 9 (0.9) | 24 (2.5) | 25 (2.5) |
| Grade 1 | 9 (0.9) | 24 (2.5) | 24 (2.4) |
| Grade 2 | 0 | 0 | 1 (0.1) |
| Erythema | 2 (0.2) | 2 (0.2) | 4 (0.4) |
| Grade 1 | 2 (0.2) | 2 (0.2) | 3 (0.3) |
| Grade 2 | 0 | 0 | 1 (0.1) |
| Bruising | 2 (0.2) | 8 (0.8) | 3 (0.3) |
| Grade 1 | 2 (0.2) | 8 (0.8) | 2 (0.2) |
| Grade 2 | 0 | 0 | 1 (0.1) |
| Pruritis | 4 (0.4) | 6 (0.6) | 6 (0.6) |
| Grade 1 | 4 (0.4) | 4 (0.4) | 6 (0.6) |
| Grade 2 | 0 | 2 (0.2) | 0 |

A participant with multiple reactions was counted only once for that reaction, and at the maximum toxicity grade.

**Supplementary Table 4.** Local Injection-Site Reactions From the Study Intervention Diary Card (Safety Set)

|  | Placebo (n = 983)<br>n (%) | VIR-2482 450 mg<br>(n = 981)<br>n (%) | VIR-2482 1200<br>mg (n = 992)<br>n (%) |
| --- | --- | --- | --- |
| Any injection-site reaction | 272 (27.7) | 292 (29.8) | 293 (29.5) |
| Grade 1 | 227 (23.1) | 250 (25.5) | 248 (25.0) |
| Grade 2 | 38 (3.9) | 35 (3.6) | 39 (3.9) |
| Grade 3 | 7 (0.7) | 7 (0.7) | 6 (0.6) |
| Pain | 260 (26.5) | 279 (28.4) | 277 (27.9) |
| Grade 1 | 222 (22.6) | 240 (24.5) | 237 (23.9) |
| Grade 2 | 31 (3.2) | 32 (3.3) | 34 (3.4) |
| Grade 3 | 7 (0.7) | 7 (0.7) | 6 (0.6) |
| Swelling | 2 (0.2) | 10 (1.0) | 13 (1.3) |
| Grade 1 | 1 (0.1) | 8 (0.8) | 9 (0.9) |
| Grade 2 | 1 (0.1) | 2 (0.2) | 4 (0.4) |
| Erythema | 5 (0.5) | 7 (0.7) | 10 (1.0) |
| Grade 1 | 4 (0.4) | 6 (0.6) | 10 (1.0) |
| Grade 2 | 1 (0.1) | 1 (0.1) | 0 |
| Bruising | 21 (2.1) | 17 (1.7) | 20 (2.0) |
| Grade 1 | 16 (1.6) | 16 (1.6) | 17 (1.7) |
| Grade 2 | 5 (0.5) | 1 (0.1) | 3 (0.3) |
| Induration | 3 (0.3) | 8 (0.8) | 12 (1.2) |
| Grade 1 | 2 (0.2) | 5 (0.5) | 11 (1.1) |
| Grade 2 | 1 (0.1) | 3 (0.3) | 1 (0.1) |

A participant with multiple reactions was counted only once for that reaction, and at the maximum toxicity grade.

**Supplemental Table 5.** Systemic Reactions From the Study Intervention Diary Card by Symptom, Overall, and by Maximum Severity (Safety Set)

|  | Placebo<br>(n = 983)<br>n (%) | VIR-2482 450 mg<br>(n = 981)<br>n (%) | VIR-2482 1200 mg<br>(n = 992)<br>n (%) |
| --- | --- | --- | --- |
| Any systemic reaction | 268 (27.3) | 265 (27.0) | 283 (28.5) |
| Mild | 209 (21.3) | 210 (21.4) | 216 (21.8) |
| Moderate | 53 (5.4) | 42 (4.3) | 52 (5.2) |
| Severe | 6 (0.6) | 13 (1.3) | 15 (1.5) |
| Potentially life-threatening | 0 | 0 | 0 |
| Headache | 175 (17.8) | 160 (16.3) | 168 (16.9) |
| Mild | 138 (14.0) | 125 (12.7) | 124 (12.5) |
| Moderate | 34 (3.5) | 27 (2.8) | 34 (3.4) |
| Severe | 3 (0.3) | 8 (0.8) | 10 (1.0) |
| Myalgia | 149 (15.2) | 135 (13.8) | 169 (17.0) |
| Mild | 130 (13.2) | 121 (12.3) | 148 (14.9) |
| Moderate | 17 (1.7) | 13 (1.3) | 17 (1.7) |
| Severe | 2 (0.2) | 1 (0.1) | 4 (0.4) |
| Malaise | 123 (12.5) | 117 (11.9) | 130 (13.1) |
| Mild | 101 (10.3) | 96 (9.8) | 98 (9.9) |
| Moderate | 20 (2.0) | 18 (1.8) | 28 (2.8) |
| Severe | 2 (0.2) | 3 (0.3) | 4 (0.4) |
| Shivering | 43 (4.4) | 44 (4.5) | 60 (6.1) |
| Mild | 38 (3.9) | 33 (3.4) | 43 (4.3) |
| Moderate | 4 (0.4) | 10 (1.0) | 13 (1.3) |
| Severe | 1 (0.1) | 1 (0.1) | 4 (0.4) |
| Fever<sup>a</sup> | 13 (1.3) | 11 (1.1) | 11 (1.1) |
| Mild | 7 (0.7) | 6 (0.6) | 8 (0.8) |
| Moderate | 5 (0.5) | 3 (0.3) | 2 (0.2) |
| Severe | 1 (0.1) | 2 (0.2) | 1 (0.1) |
| Potentially life-threatening | 0 | 0 | 0 |

A participant with multiple reactions was counted only once for that reaction, and at the maximum grade.
<sup>a</sup>Fever was graded using the Toxicity Grading Scale for Healthy Adult and Adolescent Volunteers Enrolled in Preventative Vaccine Clinical Trials.

**Supplementary Table 6.** Influenza A Subtypes for Participants With Protocol-Defined ILI With RT-PCR Confirmed Influenza A Without Concomitant Coinfection (Virology Analysis Set)

|  | Placebo<br>(n = 593) | VIR-2482<br>450 mg<br>(n = 579) | VIR-2482<br>1200 mg<br>(n = 574) |
| --- | --- | --- | --- |
| No. (%) participants<sup>a</sup> | 25 (4.2) | 24 (4.2) | 21 (3.7) |
| No. of protocol-defined ILI with RT-PCR-confirmed influenza A (without coinfection), N1 | 25 | 25 | 21 |
| H1 pdm09, no. (%) | 9 (36.0) | 4 (16.0) | 5 (23.8) |
| H3, no. (%) | 14 (56.0) | 18 (72.0) | 12 (57.1) |
| Unknown, no. (%) | 2 (8.0) | 3 (12.0) | 4 (19.1) |

ILI, influenza-like illness; RT-PCR, reverse transcriptase-polymerase chain reaction.
<sup>a</sup>If a participant had multiple protocol-defined ILI with RT-PCR–confirmed influenza A without coinfection, all (N1) were included in the summary. Percentages for influenza A subtypes are based on N1.

**Supplementary Table 7:** Sequence and Phenotypic Data Availability for Participants With RT-PCR Confirmed Influenza A Virus (Safety Set)

| Study Population | Placebo | VIR-2482<br>450 mg | VIR-2482<br>1200 mg | Total |
| --- | --- | --- | --- | --- |
|  | n/N<sup>a</sup> (%) |  |  |  |
| Participants with HA<br>sequence data available | 14/29 (48.3) | 18/30 (60.0) | 12/24 (50.0) | 44/83 (53.0) |
| Participants with phenotypic<br>data available | 15/29 (51.7) | 17/30 (56.7) | 10/24 (41.7) | 42/83 (50.6) |

RT-PCR, reverse transcriptase-polymerase chain reaction.
<sup>a</sup>n is number of participants with sequence or phenotypic data available; N is number of
participants with RT-PCR confirmed influenza A in the safety set.

**Supplementary Table 8:** Serum PK Parameters (PK Analysis Set) extravascular administration point.

| PK parameter (unit) | VIR-2482 450 mg |  |  | VIR-2482 1200 mg |  |  |
| --- | --- | --- | --- | --- | --- | --- |
|  | Thigh | Buttock | Total | Thigh | Buttock | Total |
| C<sub>max</sub> (µg/mL) |  |  |  |  |  |  |
| n | 636 | 189 | 826 | 657 | 172 | 829 |
| Geometric mean | 55.6 | 30.9 | 48.6 | 128 | 64.8 | 111 |
| Geometric CV% | 39.2 | 65.5 | 53.6 | 33.7 | 67.8 | 52.1 |
| C<sub>180</sub> (µg/mL) |  |  |  |  |  |  |
| n | 636 | 189 | 826 | 657 | 172 | 829 |
| Geometric mean | 6.1 | 4.2 | 5.6 | 14 | 8.3 | 12.6 |
| Geometric CV% | 124.8 | 70.7 | 114.8 | 130.6 | 93.3 | 127.4 |
| AUC<sub>0-180</sub> (d*µg/mL) |  |  |  |  |  |  |
| n | 636 | 189 | 826 | 657 | 172 | 829 |
| Geometric mean | 3960 | 2430 | 3540 | 9140 | 5030 | 8070 |
| Geometric CV% | 39.9 | 57.1 | 49.6 | 36.3 | 58.9 | 49.4 |
| AUC<sub>∞</sub> (d*µg/mL) |  |  |  |  |  |  |
| n | 240 | 82 | 322 | 253 | 71 | 324 |
| Geometric mean | 3970 | 2760 | 3620 | 9070 | 5830 | 8230 |
| Geometric CV% | 45.3 | 58.1 | 51.8 | 46.9 | 64.3 | 55.0 |
| CL/F (mL/d) |  |  |  |  |  |  |
| n | 240 | 82 | 322 | 253 | 71 | 324 |
| Geometric mean | 113 | 163 | 124 | 132 | 206 | 146 |
| Geometric CV% | 45.3 | 58.1 | 51.8 | 46.9 | 64.3 | 55.0 |
| $V_z/F$ (L) | | | | | | |
| n | 240 | 82 | 322 | 253 | 71 | 324 |
| Geometric mean | 7.9 | 12.8 | 9.0 | 9.2 | 15.7 | 10.3 |
| Geometric CV% | 34.7 | 55.6 | 46.6 | 30.1 | 61.0 | 45.0 |
| $T_{max}$ (d) | | | | | | |
| n | 636 | 189 | 826 | 657 | 172 | 829 |
| Median | 7.0 | 7.2 | 7.0 | 6.9 | 7.8 | 7.0 |
| Min, max | 2.8, 40.9 | 4.1, 35.8 | 2.8, 40.9 | 1.9, 57.9 | 4.9, 71.1 | 1.9, 71.1 |
| $T_{1/2}$ (d) | | | | | | |
| n | 249 | 85 | 334 | 272 | 71 | 343 |
| Median | 55.1 | 56.7 | 55.4 | 54.5 | 55.7 | 54.7 |
| Min, max | 15.5, 76.7 | 26.1, 76.8 | 15.5, 76.8 | 14, 79 | 15.3, 77 | 14, 79 |

$AUC_{\infty}$ , area under the concentration-time curve extrapolated from time zero to time infinity; $AUC_{0-180}$ , partial area under the concentration-time curve from time zero to day 180; CL/F, total body clearance following extravascular administration; $C_{max}$ , maximum observed serum concentration; $C_{180}$ , predicted serum concentration at day 180; CV, coefficient of variation; PK, pharmacokinetics; $T_{max}$ , time at which maximum serum concentration was observed; $T_{1/2}$ , terminal elimination half-life; $V_z/F$ , apparent volume of distribution during the terminal elimination phase, following extravascular administration point.

**Supplementary Table 9:** PK and Virology Data From Protocol-Defined ILI Cases.

| Location | Date of dose | Date of protocol-defined ILI | Days elapsed from dose to protocol-defined ILI | VIR-2482 serum concentration (µg/mL) | VIR-2482 NPS concentration (µg/mL) | Influenza A subtype |
| --- | --- | --- | --- | --- | --- | --- |
| Buttock 450 mg | 2022-11-01 | 2022-11-24 | 24 | 18.0 | 0.6 | H3 |
|  | 2022-11-07 | 2022-11-26 | 20 | 37.5 | NC | H3 |
|  | 2022-10-28 | 2022-11-03 | 7 | 18.9 | NC | H1pdm09 |
| Buttock 1200 mg | 2022-11-04 | 2022-12-04 | 31 | 15.9 | 2.1 | H3 |
|  | 2022-11-17 | 2022-11-30 | 14 | – | NC | Unknown |
| Thigh 450 mg | 2022-11-21 | 2022-12-19 | 29 | 32.4 | NC | H3 |
|  | 2022-11-11 | 2022-12-01 | 21 | 46.1 | NC | Unknown |
|  | 2022-11-21 | 2022-12-05 | 15 | 45.0 | NC | H3 |
|  | 2022-11-02 | 2022-12-01 | 30 | 15.9 | NC | H1pdm09 |
|  | 2022-11-16 | 2022-11-28 | 13 | 70.4 | – | Unknown |
|  | 2022-11-10 | 2022-11-21 | 12 | 65.1 | NC | H3 |
|  | 2022-11-08 | 2022-12-28 | 51 | 35.8 | NC | H3 |
|  | 2022-11-11 | 2022-11-23 | 13 | 37.8 | NC | H3 |
|  | 2022-11-28 | 2023-01-12 | 46 | BQL | NC | H1pdm09 |
|  | 2022-11-15 | 2022-12-13 | 29 | 47.0 | NC | H3 |
|  | 2022-10-28 | 2022-12-01 | 35 | 53.4 | NC | H3 |
|  | 2022-11-28 | 2023-02-13 | 78 | 34.6 | NC | H3 |
|  | 2022-11-28 | 2023-02-13 | 78 | 25.1 | NC | H3 |
|  | 2022-11-18 | 2022-11-21 | 4 | 74.6 | 2.8 | H3 |
|  | 2022-11-28 | 2022-12-12 | 15 | 80.8 | NC | H3 |
|  | 2022-10-26 | 2022-11-23 | 29 | 26.3 | NC | H3 |
|  | 2022-11-09 | 2022-12-15 | 37 | 28.0 | 2.5 | H3 |
|  | 2022-11-10 | 2022-11-24 | 15 | 46.7 | 1.0 | H3 |
|  | 2022-11-08 | 2022-12-21 | 44 | 27.8 | – | Unknown |
|  | 2022-11-16 | 2022-12-07 | 22 | 58.1 | – | H1pdm09 |
|  | 2022-11-16 | 2022-11-24 | 9 | 66.8 | NC | H3 |
| Thigh 1200 mg | 2022-11-16 | 2022-11-19 | 4 | 205.6 | NC | H3 |
|  | 2022-11-10 | 2022-11-13 | 4 | 102.5 | 0.7 | H3 |
|  | 2022-11-17 | 2022-12-05 | 19 | 98.2 | NC | H3 |
|  | 2022-11-28 | 2022-12-27 | 30 | 69.7 | 2.4 | H3 |
|  | 2022-12-01 | 2022-12-08 | 8 | 169.2 | 0.9 | H3 |
|  | 2022-12-01 | 2022-12-05 | 5 | 119.0 | NC | H3 |
|  | 2022-11-28 | 2022-12-02 | 5 | 173.5 | 4.2 | H1pdm09 |
|  | 2022-11-22 | 2022-12-10 | 19 | 170.4 | NC | H3 |
|  | 2022-11-07 | 2022-11-28 | 22 | 119.9 | NC | H3 |
|  | 2022-10-28 | 2022-11-10 | 14 | 122.0 | – | Unknown |
|  | 2022-10-19 | 2023-03-15 | 148 | 33.4 | NC | H1pdm09 |
|  | 2022-12-05 | 2022-12-15 | 11 | 101.5 | NC | H1pdm09 |
|  | 2022-11-07 | 2022-12-04 | 28 | 103.9 | NC | H1pdm09 |
|  | 2022-12-05 | 2022-12-05 | 1 | 43.3 | NC | H3 |
|  | 2022-10-26 | 2022-12-12 | 48 | 57.8 | – | Unknown |
|  | 2022-11-22 | 2022-12-05 | 14 | 94.1 | 2.6 | H3 |
|  | 2022-11-15 | 2022-12-01 | 17 | 83.6 | 3.0 | H1pdm09 |
|  | 2022-12-01 | 2022-12-28 | 28 | 116.9 | — | Unknown |
|  | 2022-12-02 | 2022-12-12 | 11 | 126.3 | 0.8 | H3 |

BQL, below the quantification limit; PK, pharmacokinetics; ILI, influenza-like illness; NC, not calculable; NPS, nasopharyngeal swab.

**Supplementary Figure 1.**
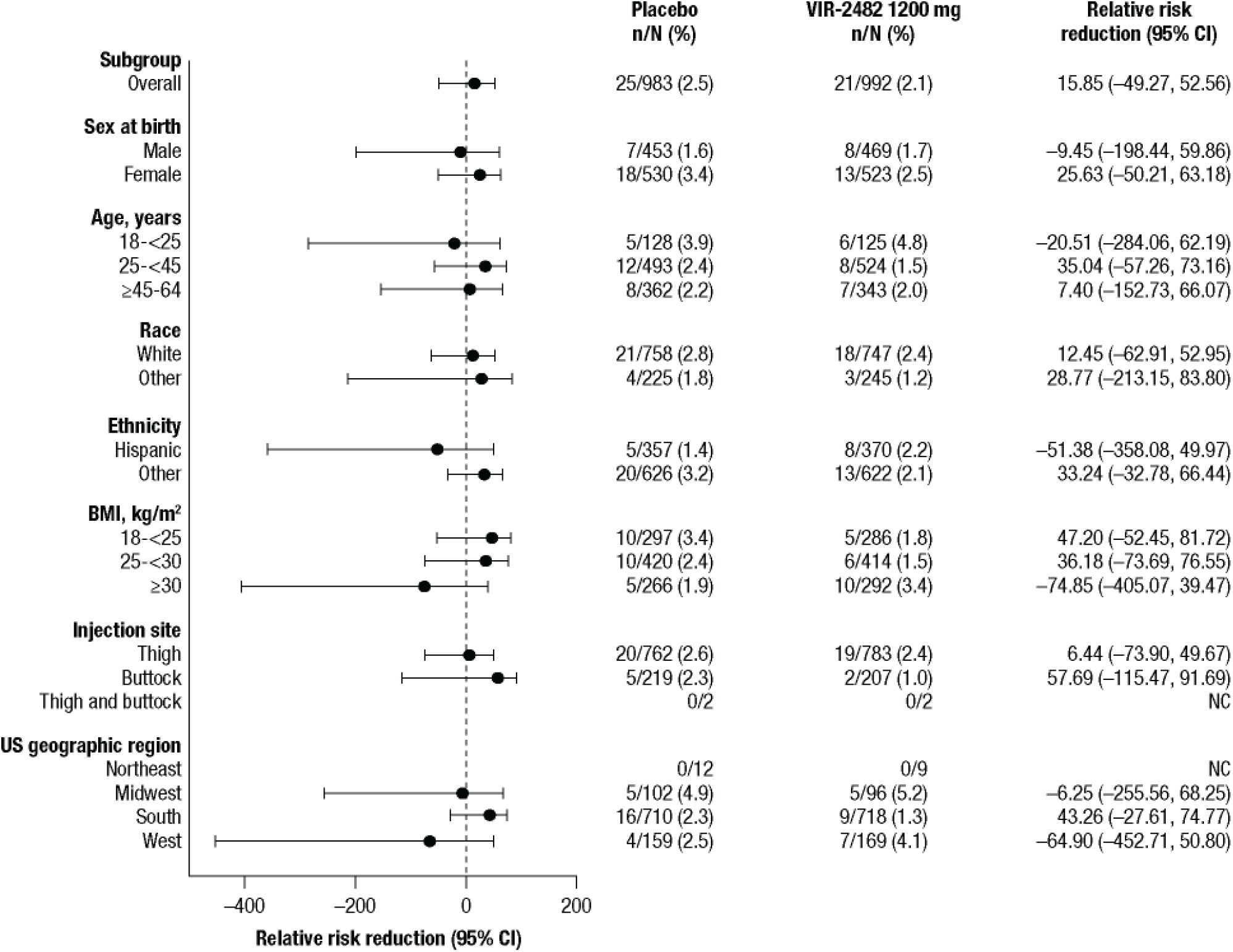
Subgroup Analyses for the Occurrence of Protocol-Defined Influenza-Like Illness With RT-PCR Confirmed Influenza A in Participants Receiving VIR-2482 1200 mg. BMI, body mass index; NC, not calculable; RT-PCR, reverse transcriptase-polymerase chain reaction.

**Supplementary Figure 2.**
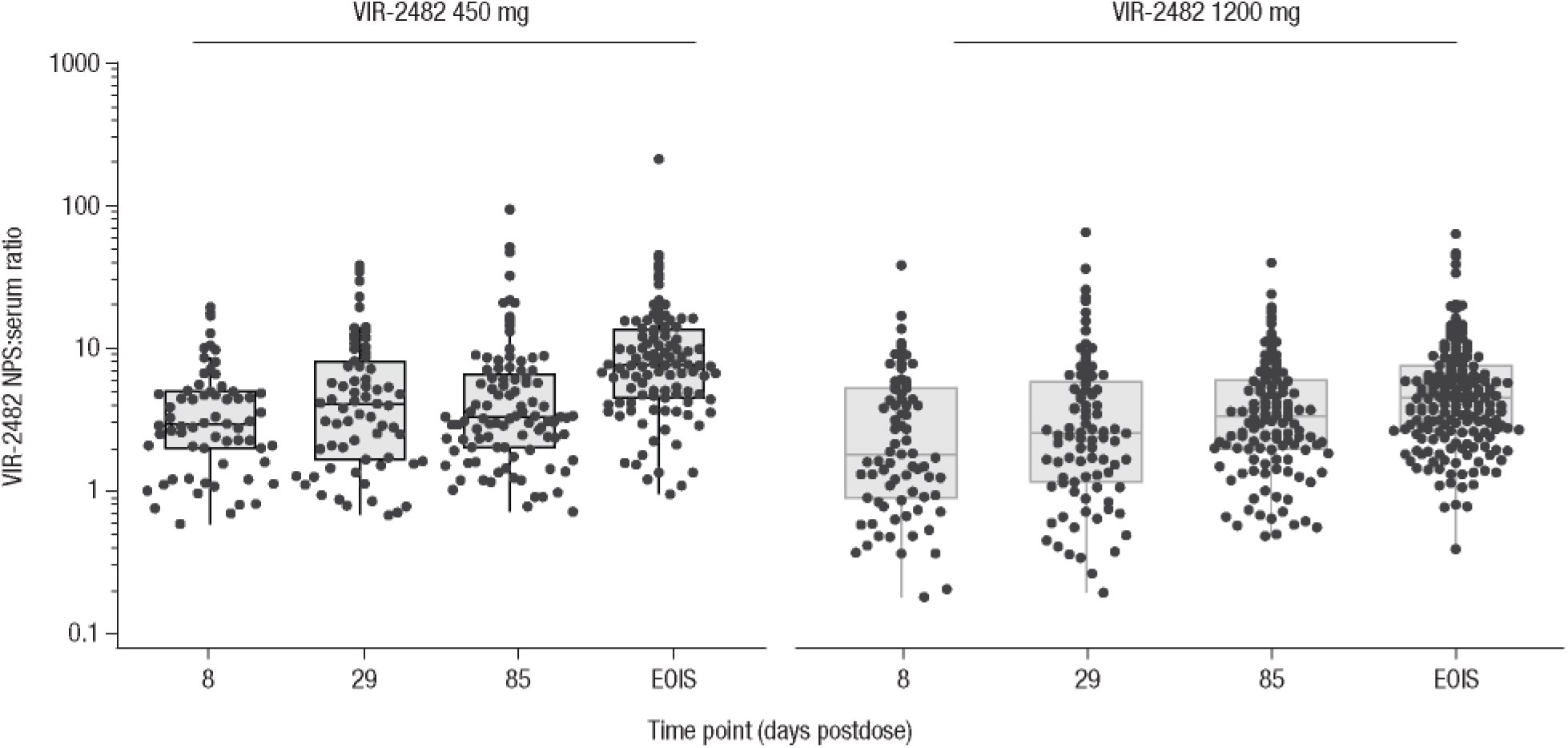
VIR-2482 NPS:Serum Ratios by Dose and Timepoint (PK Analysis Set) EOIS, end of influenza season; NPS, nasopharyngeal swab; PK, pharmacokinetics. The ratio of VIR-2482 in NPS samples versus serum is calculated as 100 x (VIR-2482 NPS concentration/VIR-2482 serum concentration.57

